# AI-Driven Analysis of Device Duration in Anesthesiology

**DOI:** 10.64898/2026.09.25.26364018

**Authors:** Arnab Sutar, Brandon Tabman, Huda Fatima, Payam Norouzzadeh, Yuri Martins, Mostafa Yazdi, Bahareh Rahmani

## Abstract

Lines, drains, and airway devices are commonly used in surgical and inpatient care to support treatment and patient monitoring. While these devices are essential, prolonged device retention is associated with complications such as infection, tissue injury, thrombosis, and extended hospital stay. Understanding how long devices remain in place and how duration varies across device types is important for improving patient safety and guiding device management practices. This study analyzes device duration patterns using Patient Lines, Drains, and Airways (LDA) data containing information on device type, placement time, and removal time across multiple patient encounters. Exploratory data analysis and machine learning approaches are used to examine duration distributions, distinguish short and prolonged device retention, and identify structural patterns across device categories. The findings indicate notable variation in device duration patterns across categories, with identifiable differences in device retention behavior across device groups. This study highlights the value of observational electronic health record data for characterizing device exposure patterns and provides a foundation for future datadriven device management and risk assessment.

## 1. Introduction

Lines, drains, and airway devices are essential components of modern surgical and inpatient care. These devices include vascular access lines, surgical drains, chest tubes, endotracheal tubes, and tracheostomies. They play a critical role in patient monitoring, medication delivery, and respiratory support. However, prolonged retention of these devices is associated with adverse outcomes such as catheter-related infections, surgical site infections, airway complications, thrombosis, and delayed recovery [1–5].

Device-related complications have been widely studied, particularly for individual device types. Researchers investigating vascular access devices have demonstrated that catheter use and prolonged dwell time are associated with an increased risk of bloodstream infection and other complications, informing clinical guidelines that emphasize appropriate catheter selection and timely device removal [1–4]. Similar concerns exist for central venous access devices, for which complications include infection, thrombosis, and mechanical complications [5]. Surgical drains have also been extensively studied, with evidence suggesting that drain use and prolonged drainage may influence surgical site infection and postoperative outcomes [6–9]. Studies of airway devices demonstrate that prolonged mechanical ventilation and delayed tracheostomy are associated with increased risk of complications and longer hospital stays [10–13].

Early research on device duration focused primarily on device-specific guidelines and clinical recommendations. While these studies provide important insights, they often examine devices in isolation and do not assess duration patterns across multiple device categories. With the increasing use of electronic health record systems, detailed documentation of clinical events and device-related information has become available, allowing for large-scale analysis of healthcare processes and patient-level exposure patterns across diverse populations [14–16].

Researchers analyzing observational healthcare data have noted several analytical challenges. Device usage patterns vary widely across patients and clinical contexts, and duration distributions may overlap substantially across device types. Traditional statistical approaches may therefore have limited ability to fully explain or predict complex patterns in healthcare data. In response, researchers have increasingly applied machine learning methods to large observational healthcare datasets because these approaches can capture nonlinear relationships, complex interactions, and patterns that may be difficult to model using conventional statistical techniques [17–20].

In related areas of healthcare data analysis, researchers have used machine learning approaches to explore structure in high-dimensional observational data and to support risk stratification, prediction, and clinical decision-making [17–20]. These approaches can help identify patterns that may not be readily apparent through conventional analyses. However, evidence regarding the application of machine learning specifically to device duration and retention patterns across multiple lines, drains, and airway categories remains limited.

In this study, we analyze Patient Lines, Drains, and Airways (LDA) data to examine duration patterns across twelve categories of devices. We apply exploratory data analysis and classification models to characterize device duration behavior and evaluate the extent to which device category alone explains prolonged retention. Rather than prioritizing predictive accuracy, this study focuses on understanding structural patterns and limitations inherent in observational device duration data. The findings provide insight into device exposure behavior and highlight opportunities for future data-driven approaches to device management, anesthesiology and perioperative care, and patient safety.

## 2. Data Description

We used the Patient Lines, Drains, and Airways (LDA) dataset from the University of California, Irvine repository. The dataset contains electronic health record documentation of device use during inpatient encounters, including device name, placement time, removal time, and duration of use in days. Multiple devices may be recorded within a single patient encounter. Device names were mapped to standardized line group categories representing major device types such as vascular access lines, drains, airway devices, and related device groups.

Before analysis, we removed records with missing device group or duration information. Device duration was calculated from placement and removal timestamps and expressed in days. The cleaned dataset was used for all duration summaries, modeling, and pattern analysis presented in this study.

## 3. Methodology

We analyzed Patient Lines, Drains, and Airways data to examine how device duration varies across standardized line group categories. Device duration was measured in days using documented placement and removal times. We assessed whether line group alone explains differences in retention behavior using classification approaches. Visual summaries and tables were generated to illustrate frequency, duration patterns, and structural variability across device categories.

### 3.1 Predictive Modeling of Device Duration

We constructed binary classification models to distinguish lower versus higher device duration based on the median duration threshold. Line group was encoded using one-hot encoding and used as the sole predictor. XGBoost, Support Vector Machine, and Random Forest classifiers were trained. Model performance was evaluated using overall classification accuracy on a held-out test set.

## Results

### 4.1 Distribution of Device Records Across Line Groups

The distribution of device records varied substantially across line groups. Wound-related devices were most frequent, followed by PIV lines and drains. In contrast, nasogastric/orogastric tubes and extravasation-related devices were rare. This imbalance reflects substantial differences in routine clinical utilization across device types (Table 1).

**Table 1.** Counts of records by line group.

| Line Group Name | Num records |
| --- | --- |
| Wound | 63457 |
| PIV Line | 57241 |
| Drain | 41874 |
| Urinary Drainage | 31619 |
| CVC Line | 9773 |
| ART Line | 5342 |
| Airway | 5269 |
| PICC Line | 4241 |
| LineType | 2986 |
| Epidural Line | 906 |
| Nasogastric/Orogastric tube | 89 |
| Extravasation | 51 |

To visually illustrate these differences, we present the distribution graphically in figure 1.

**Figure 1.**
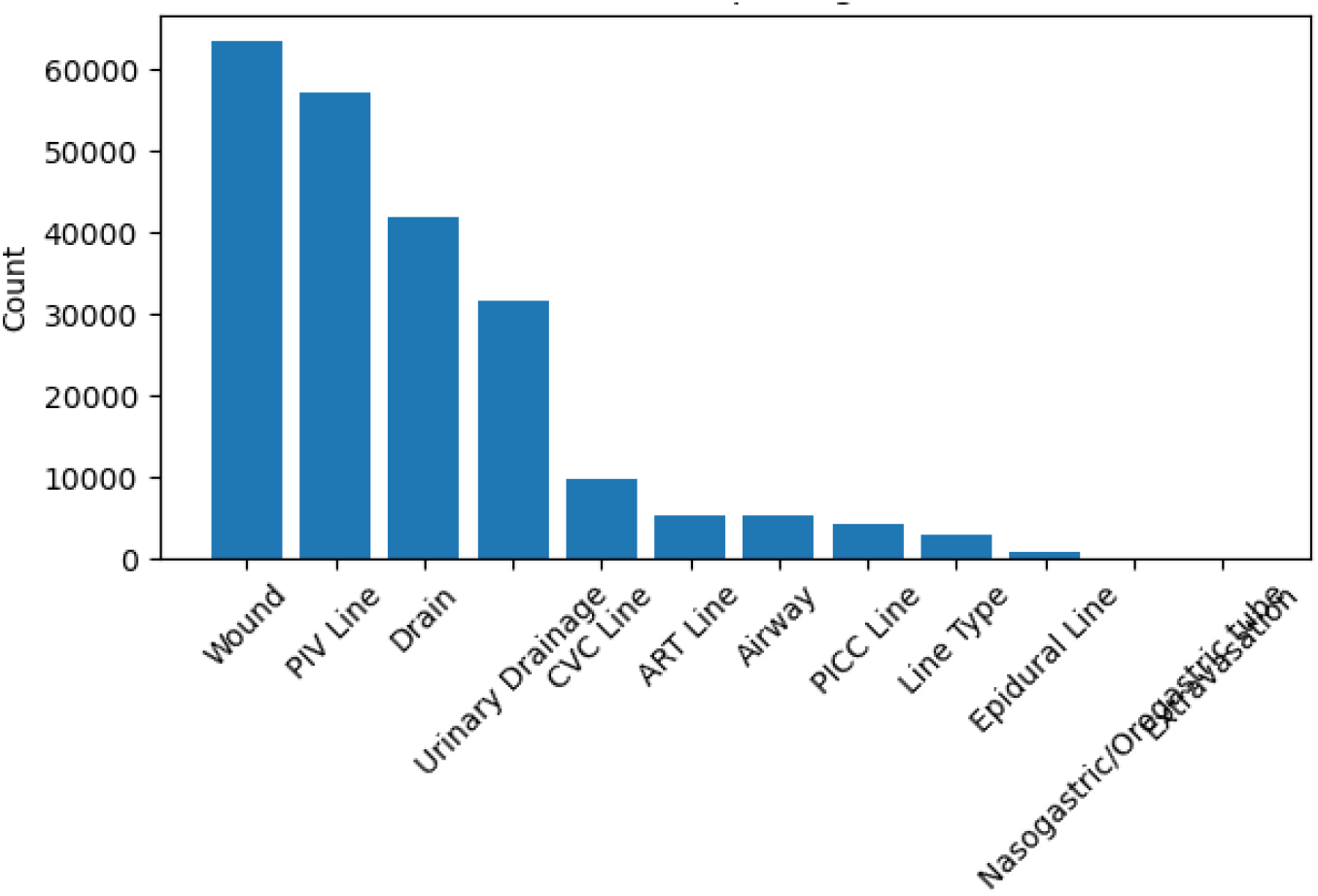
Counts of Device Records by Line Group.

### 4.2 Total Duration by Line Group

While record counts indicate frequency of use, total cumulative duration reflects overall exposure burden. Wound-related devices account for the highest total duration, consistent with their high utilization. CVC lines, line type devices, drains, and PICC lines also contribute substantially, whereas nasogastric/orogastric tubes, extravasation-related devices, and epidural lines contribute minimally. This contrast highlights differences between device frequency and retention intensity across categories shown in figure 2.

**Figure 2.**
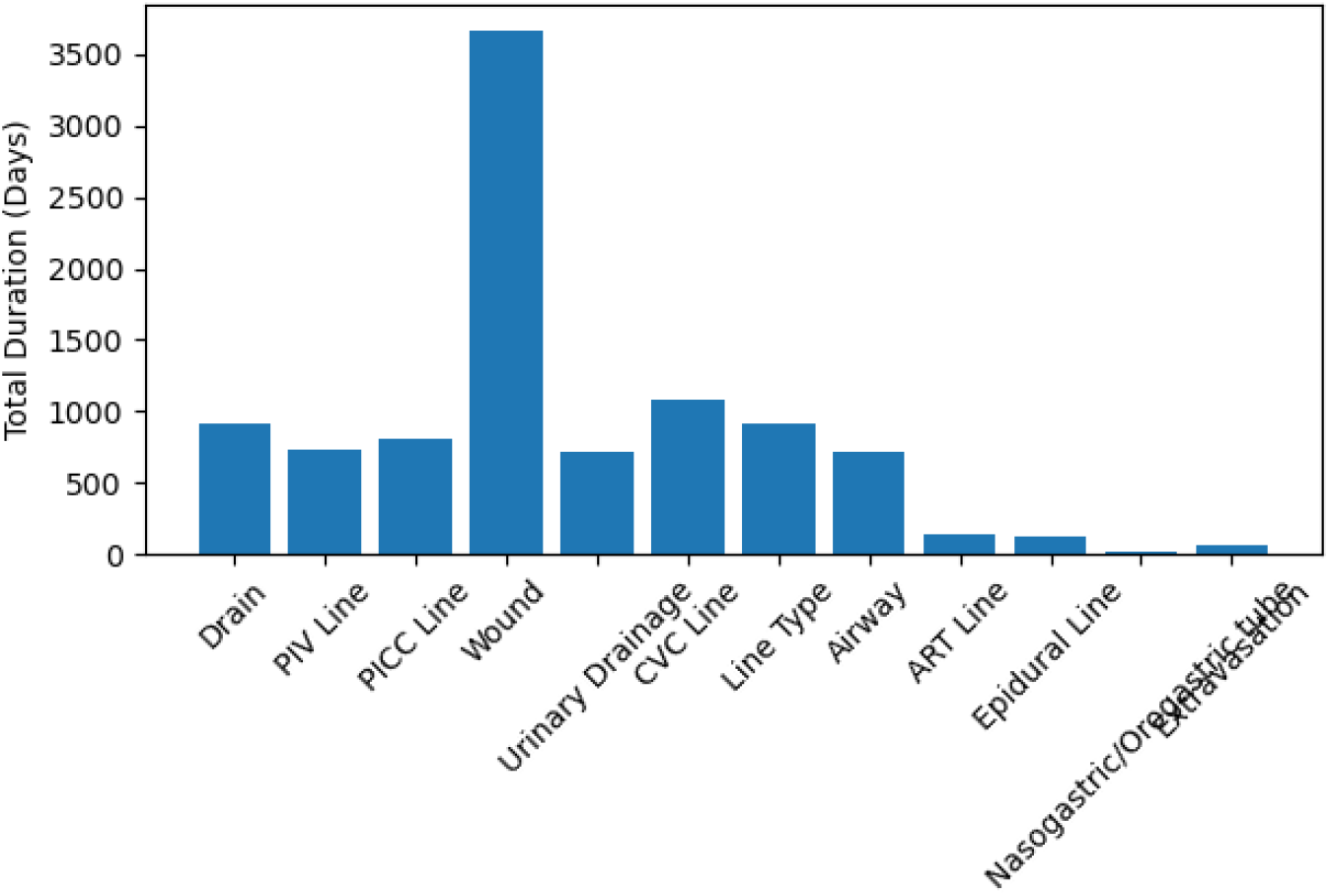
Total Cumulative Device Duration by Line Group.

### 4.3 Binary Classification of Device Duration

Binary classification models were developed to distinguish lower versus higher device duration categories using line group as the sole predictor. XGBoost, Support Vector Machine (SVM), and Random Forest classifiers were trained and evaluated using overall classification accuracy on a held-out test dataset. All three models demonstrated highly similar performance, each achieving an accuracy of approximately 61.7% in distinguishing lower versus higher device duration categories.

Despite differences in algorithmic structure, the similarity in model performance can be attributed primarily to the structure of the input data rather than limitations of the modeling approach. The models were trained using a single predictor variable, *Line_Group_Name*, which was encoded using one-hot encoding. Consequently, the feature space available to the algorithms consisted only of categorical indicators representing the different line groups.

With this restricted feature set, the models primarily learned the relationship between line group category and the binary duration outcome. Because the underlying information available to each algorithm was essentially the same and relatively simple, the models converged toward similar decision boundaries and produced comparable predictions on the test data. Such consistency across modeling approaches is common in machine learning studies where the available predictors provide limited variability. These results indicate that line group category captures meaningful structural patterns related to device duration while also highlighting the limitations of relying on a single categorical predictor.

**Table 2.**
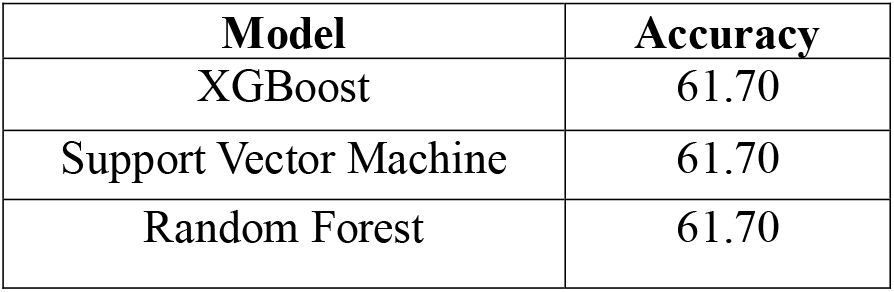
Binary Classification Performance.

### 4.3.1 Receiver Operating Characteristic (ROC) Curve Analysis

Receiver Operating Characteristic (ROC) curves were generated for XGBoost, Support Vector Machine, and Random Forest to evaluate classification performance across probability thresholds. As illustrated in Figures 3, all three models demonstrated nearly identical ROC curves, with AUC values of approximately 0.653. The close alignment of the curves indicates consistent discrimination between lower and higher device duration categories across the different modeling approaches. Because all models were trained using the same predictor variable, *Line_Group_Name*, the underlying information available to each algorithm was essentially identical, resulting in closely aligned probability estimates and comparable ROC–AUC values shown in figure 3.

**Figure 3.**
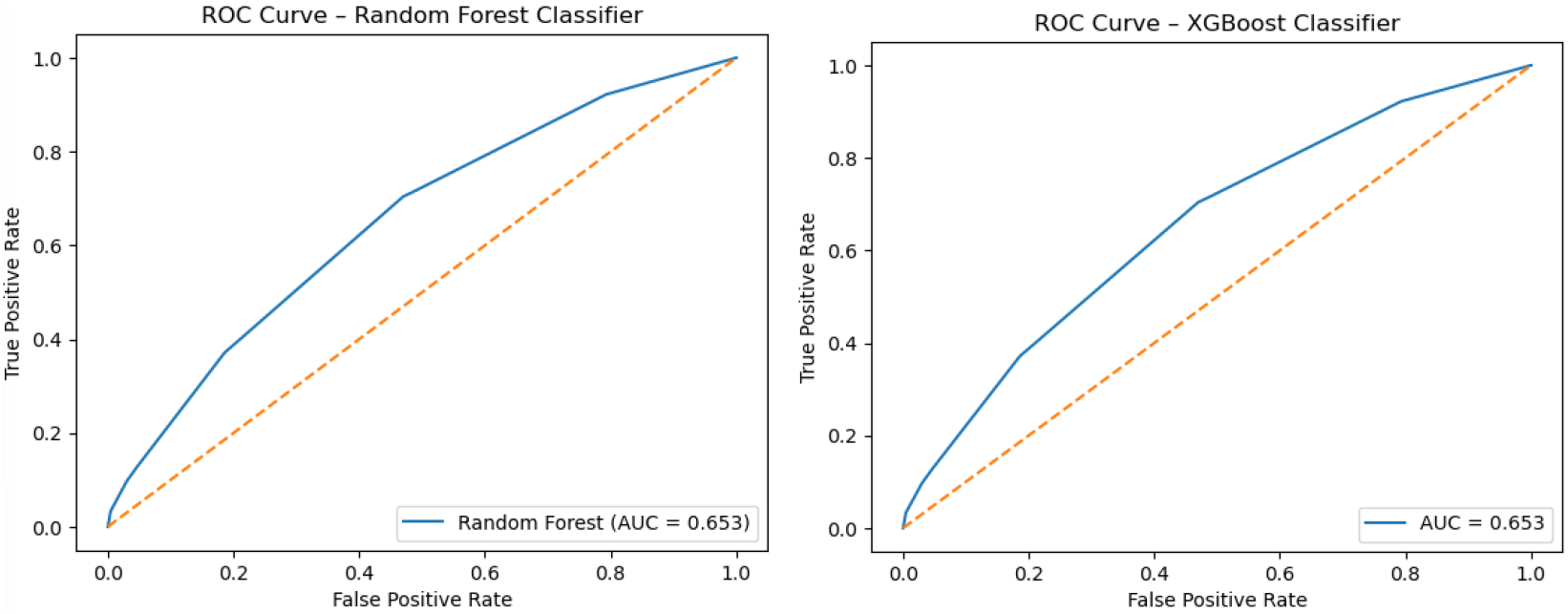
ROC Curve Comparison Across Classification Models.

## 5. Discussion

This study analyzed device duration patterns using Patient Lines, Drains, and Airways data derived from observational electronic health record documentation to assess whether standardized line group categories explain differences in retention behavior. Substantial variation was observed in both frequency of use and cumulative retention across device types. Wound-related devices and PIV lines were most frequently documented, yet frequency did not uniformly translate into proportional exposure burden, indicating differences in retention behavior across categories.

Binary classification models demonstrated consistent performance across XGBoost, Support Vector Machine, and Random Forest algorithms shown in figure 4. All three models achieved an overall accuracy of approximately 61.7%, with AUC values of 0.653. The near-identical ROC curves further confirm that model discrimination was stable and reproducible across different supervised learning approaches. This consistency reflects the fact that the models were trained using the same categorical predictor variable, which limited the variability available to the algorithms while still capturing meaningful structural patterns associated with device duration.

**Figure 4.**
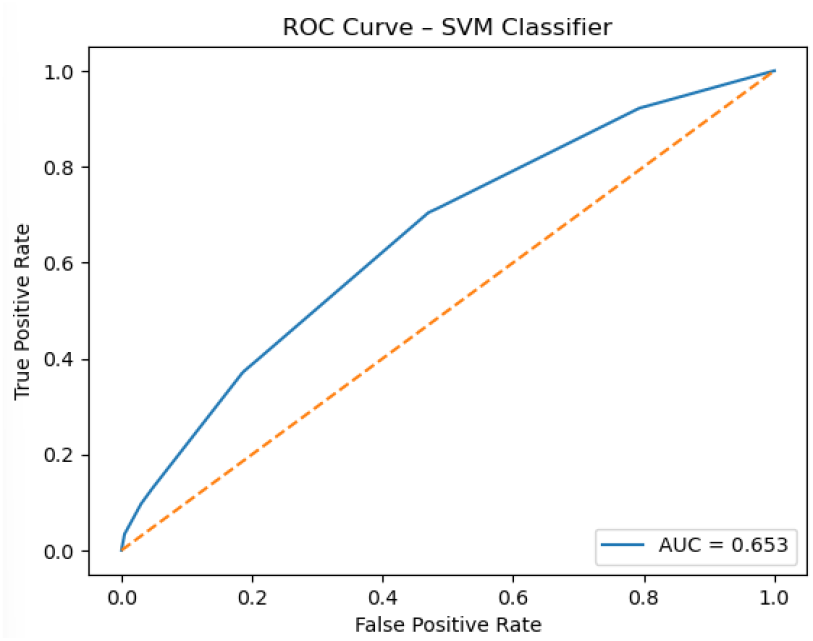
ROC Curve – SVM Classifier.

Overall, line group category explains important patterns in retention and reveals structured variability in duration behavior across line groups. These findings demonstrate the value of supervised classification approaches to better characterize device exposure patterns in observational healthcare data.

## 6. Conclusion

This study characterized device duration and retention patterns across twelve categories of lines, drains, and airway devices using electronic health record data. The analysis demonstrated substantial variation in device frequency and cumulative duration across device groups, highlighting meaningful differences in device exposure patterns.

The main findings can be summarized as follows:

- **Substantial variation across device types:** Device frequency and cumulative duration differed considerably among the twelve line groups, indicating distinct patterns of device utilization and retention.
- **Moderate predictive signal from device category:** Using line group as the sole predictor, XGBoost, Support Vector Machine, and Random Forest achieved approximately **61.7% accuracy** and an **AUC of 0.653**.
- **Consistent performance across models:** The similar performance of the three algorithms suggests that the predictive signal was primarily driven by the underlying categorical structure of device type rather than by differences in modeling methodology.
- **Device category alone is insufficient:** Although line group provides meaningful information about duration, its moderate predictive performance indicates that device retention is influenced by factors beyond device type.
- **Foundation for future clinical modeling:** Incorporating patient characteristics, clinical conditions, procedural factors, device-specific features, and perioperative variables may improve prediction of prolonged device retention.

Overall, this study provides a data-driven characterization of lines, drains, and airway device duration patterns and establishes a baseline for future machine learning research. These findings may support the development of more comprehensive approaches to device management and risk assessment in **anesthesiology, perioperative care, and inpatient medicine**, with the broader goal of improving patient safety and reducing potentially avoidable prolonged device exposure.

## Data Availability

All data produced in the present study are available upon reasonable request to the authors

## Author Contribution

Arnab Sutar, Write the Original Paper, Programming, Machine Learning Modeling

Brandon Tabman, Data Curation, Visualization, Data Analysis, Programming

Huda Fatima, Literature Review, Data Curation, Data Analysis

Payam Norouzzadeh, Methodology, Review the Paper

Yuri Martins, Supervision, Validation, Review the Paper

Mostafa Yazdi, Review the paper, Revision the Manuscript

Bahareh Rahmani, Administration, Define the Project, Methodology, Validation, Review the Paper

